# Associations between symptoms of depression and anxiety with left ventricular hypertrophy among Hispanic/Latino participants of the Hispanic Community Health Study/Study of Latinos

**DOI:** 10.64898/2026.01.15.26343818

**Authors:** Sharon N. Andrade-Bucknor, Robert A. Mesa, Christina Cordero, Neil Schneiderman, Alvin Mathew, Barry E Hurwitz, Carlos J. Rodriguez, Linda C. Gallo, Carlos E. Rosas, Sara Gonzalez, Scott D. Solomon, Susan Cheng, Martha Daviglus, Mayank M Kansal, Krista M. Perreira, Frank J. Penedo, Tali Elfassy

## Abstract

**Background:** Left ventricular hypertrophy (LVH) is a major independent risk factor for cardiovascular disease and is the leading cause of death among all U.S. groups, including Hispanic/Latino adults. LVH is commonly seen in the setting of hypertension and there is evidence that psychological factors, such as depression and anxiety, are risk factors in hypertension development.

**Objective:** To evaluate the association between symptoms of depression and trait anxiety with incident LVH among U.S. Hispanic/Latino adults.

**Methods:** The Hispanic Community Health Study/Study of Latinos is an ongoing population-based observational cohort study of Hispanic/Latino adults. Participants were examined in 2008-2011 at visit 1 (V1) and in 2014-2017 visit 2 (V2). Symptoms of depression and trait anxiety were assessed by self-reported questionnaire collection. LVH was assessed by echocardiography at V2. Multivariable Poisson regression models were used to determine the associations between symptoms of depression and trait anxiety at V1 with incident LVH at V2 among 6,612 participants after adjustment for potential confounders. All analyses accounted for the complex survey design and incorporated study weights.

**Results:** After an average follow-up of 6.0 years, the age-standardized cumulative incidence of LVH was 5.4% (95% CI: 4.9, 6.1). The cumulative incidence of LVH was higher among participants with (10.4%, 95% CI: 8.6, 12.4) compared to without elevated symptoms of depression (5.1%; 95% CI: 4.4, 5.9). Compared with the lowest trait anxiety tertile (5.2%; 95% CI: 4.3, 6.3), the cumulative incidence of LVH was higher in the highest trait anxiety tertile (9.6%; 95% CI: 8.1, 11.5). In multivariable Poisson models, each standard deviation increment in symptoms of depression was associated with a 10% greater liklihood (IDR:1.10, 95% CI: 1.00, 1.20) of LVH at V2. However, symptoms of trait anxiety at visit 1 were not independently associated with LVH at visit 2.

**Conclusion:** Greater depressive but not trait anxiety symptomatology was associated with LVH over six years. In fully adjusted models, a one-unit increase in depression symptomatology was associated with a 10% greater likelihood of LVH. Further studies are needed to examine the etiological role of negative affective psychological regulation.

## INTRODUCTION

Left ventricular hypertrophy (LVH) is defined by increased left ventricular mass, either due to increased cavity size or wall thickening, or both,^1^ and is a major independent risk factor for cardiovascular disease (CVD),^2^ the leading cause of death in the US and globally. More specifically, the presence of LVH is associated with heart failure,^3,4^ coronary artery disease,^5^ stroke,^6^ arrhythmias,^7^ and sudden cardiac death.^8^ In fact, the risk of death or nonfatal complications is two to four-fold higher in the presence of LVH, independent of other risk factors.^5,9^ Conversely, LVH regression is associated with lower cardiovascular risk, independent of BP lowering.^10–13^ Therefore, identifying and addressing causes or contributors to LVH may reduce the burden of CVD. Although the prevalence of LVH is high among all adults,^14^ findings from the Multi-Ethnic Study of Atherosclerosis (MESA) study showed that left ventricular mass index (LVMI), as an index of LVH, was more predictive of future CVD events among Hispanic/Latino adults compared with non Hispanic/Latino White adults.^15^ Yet studies^16–18^ investigating predictors of LVH among US Hispanic/Latino adults, who constitute the largest ethnic minority population,^19^ have been sparse.

LVH is typically seen in prolonged states of increased afterload which causes an initially adaptive growth response within the myocardium to maintain cardiac output. Several risk factors for the LVH development have been recognized, most commonly hypertension, however there are others including age, obesity, diabetes, hypercholesterolemia, prior myocardial infarction, aortic stenosis and other valvular heart disease, African American race (with social adversity and genetic variations playing a role)^17^, and high sodium intake (independent of arterial pressure).^20–22^ Additionally, there is evidence to support the effect of psychological factors, such as anxiety,^23–26^ and to a lesser extent, depression,^26–28^ as risk factors for the development of hypertension. However, whether symptoms of depression or trait anxiety are associated with LVH has gained little attention in the literature^29^ in the Hispanic/Latino population. This is especially relevant in the context of increasing levels of mental distress as a result of impaired emotional regulation reported in the U.S. and around the world.^30^

Among U.S. Hispanic/Latino adults, research from the Hispanic Community Health Study/Study of Latinos (HCHS/SOL) has shown that elevated levels of anxiety^31^ and depression symptomatology^32^ are associated with greater longitudinal increase in systolic blood pressure and a higher incidence of hypertension. Within this context, the goal of this work is to investigate whether symptoms of depression and trait anxiety are associated with LVH among U.S. Hispanic/Latino participants of the HCHS/SOL. We hypothesized that, as seen in other populations, symptoms of depression and anxiety would be associated with a greater likelihood of LVH.

## METHODS

### Study Population

The HCHS/SOL is an ongoing, population-based cohort study of 16,415 diverse Hispanic/Latino adults.^33,34^ Participants aged 18-74 were recruited from one of four U.S. urban communities (Bronx, NY; Chicago, IL; Miami, FL; and San Diego, CA). Participants were examined initially at visit 1 (V1) between 2008-2011 and re-examined at visit 2 (V2) approximately six years later between 2014-2017. At both visits, participants completed standardized procedures and assessments (questionnaires, medical examinations, and laboratory measurements of blood and urinalysis samples). Each recruiting center’s local institutional review board approved the study protocol, and written informed consent was obtained from all participants before the study procedures were initiated.

### Symptoms of depression and anxiety

Depressionsymptoms were ascertained from the V1 and V2 questionnaire collection. Depression symptoms were measured through the Center for Epidemiologic Studies Depression Scale-10 (CES-D 10).^35^ Prior research has validated CES-D 10 in the evaluation of depression symptoms in Hispanic/Latino adults.^36^ Participants selected from one of four responses on the 10-item scale. Depression scores ranged from 0 to 30, with a higher score indicative of greater symptomatology. Depression symptoms were modeled as z-scores. Additionally, we dichotomized depression symptoms as elevated or not and used a cutpoint of ≥10 for elevated depression symptomatology based on prior literature.^36–38^

At V1, trait anxiety was measured from questionnaire collection using the 10-item Spielberger Trait Anxiety Scale.^39^ Participants selected one of four responses on each item, with total trait anxiety scores ranging from 0 to 40. A higher score is indicative of higher trait anxiety. Trait anxiety was modeled as z-scores and also categorized as tertiles. At V2, symptoms of generalized anxiety were mesured using the Generalized Anxiety Disorder-7 Scale^40,41^ (GAD-7). Participants selected from one of four responses on each item, with total sums ranging from 0 to 21. GAD was modeled as z-scores and categorized as ‘Elevated’ or ‘Not Elevated’ using a cut-off of 10.^40^

### Left Ventricular Hypertrophy

Based on guidelines set by the American Society of Echocardiography/European Association of Echocardiography, at V2, LVH was diagnosed using sex-specific thresholds of LVMI obtained from an echocardiogram.^42–44^ For men, a LVMI > 115 g/m^2^ indicated the presence of LVH, while in women, a LVMI > 95 g/m^2^ indicated the presence of LVH. At V1, LVH was determined from an electrocardiogram (ECG). ECGs were centrally read by the HCHS/SOL Central ECG Reading Center (EPICARE), and standardized programming was used to determine the presence or absence of LVH.

### Other Variables

Participants self-reported their age (in years), demographic characteristics (sex [male or female], education [less than or at least a high school education], Hispanic/Latino background [Central American, Cuban, Dominican, Mexican, Puerto Rican, South American, or Other], income [less than or at least a $30K annual household income], health insurance status [yes or no], and lifestyle behaviors (alcohol use [current or former/never], smoking use [current or former/never], and diet score [measured via the Alternative Healthy Eating Index-2010]^45^). Follow-up time between V1 and V2 was also recorded.

### Analytic Sample

Of the 16,415 participants at V1, the analytic sample was restricted to those who had complete information on trait anxiety and depression measures at V1, those who did not have LVH from V1 ECG, and those with echocardiography at V2. r The final analytic sample consisted of 6,612 adults (**Supplemental Figure 1**). In a cross-sectional sensitivity analysis at V2, of the 11,623 V2 participants, the analytic sample was restricted to those with complete information on generalized anxiety and depression at V2, and those with non-missing echocardiography information at V2.

### Statistical Analysis

Demographic, socio-economic, behavioral, and clinical characteristics of the sample were evaluated and described. Next, the age-standardized prevalence of LVH overall and according to the tertile of trait anxiety and elevated depression symptoms was calculated, testing for differences using chi-square tests. To assess the association between anxiety and depression symptoms at V1 with LVH at V2 multivariable adjusted Poisson regression models were used with follow-up time included as an offset in all models. To assess the cross-sectional association between symptoms of generalized anxiety and depression symptoms at V2 with LVH at V2 logistic regression models were used. Model 1 was adjusted for demographic factors (age, sex, and Hispanic/Latino background group). Model 2 was adjusted for demographic and socio-economic factors (education, income, and health insurance). In sensitivity analyses, adjustments were made for potential mediators (alcohol use, smoking status, and diet score), as well as hypertension in separate models. All analyses were conducted in SUDAAN and accounted for the sampling weights and complex survey design of the HCHS/SOL study.

## RESULTS

At V1, the mean age of the population was 51.6 years (**Table 1**). More than half (53.1%) were women, 7.1% were Central American, 25.1% were Cuban, 7.2% were Dominican, 35.1% were Mexican, 17.2% were Puerto Rican, and 5.1% were South American. Approximately one third (35.1%) had less than a high school education, 63% had less than a $30K annual income, and 53.2% had health insurance. At V1, the mean trait anxiety score was 17.0 and the mean CES-D 10 score was 7.3 with 29.6% reporting elevated depression symptoms at V1..

**Table 1:** Baseline demographics among HCHS/SOL participants, 2008-2011.

| Characteristic | Mean<br>(95% CI) | Proportion<br>(95% CI) |
| --- | --- | --- |
| **Age, mean** | 51.6 (51.2, 52.0) |  |
| **Female** |  | 53.1 (51.5, 54.7) |
| **Hispanic/Latino background** |  |  |
| Central American |  | 7.1 (6.0, 8.2) |
| Cuban |  | 25.1 (21.8, 28.8) |
| Dominican |  | 7.2 (6.0, 8.6) |
| Mexican |  | 35.1 (31.9, 38.5) |
| Puerto Rican |  | 17.2 (15.4, 19.2) |
| South American |  | 5.1 (4.3, 5.9) |
| Mixed/Other |  | 3.2 (2.6, 3.9) |
| **Less than HS education** |  | 35.1 (33.2, 37.2) |
| **Income < \$30,000** | | 63.0 (60.7, 65.2) |
| **Has health insurance** |  | 53.2 (50.9, 55.6) |
| **Current alcohol use** |  | 47.1 (45.1, 49.0) |
| **Current smoker** |  | 19.7 (18.2, 21.3) |
| **Diet score (AHEI-2010)** | 49.6 (49.2, 50.0) |  |
| **Trait anxiety score** | 17.0 (16.8, 17.2) |  |
| **CES-D 10, mean** | 7.3 (7.1, 7.5) |  |
| **Elevated depression symptoms** | 29.6 (28.1, 31.2) |  |

Abbreviations: CES-D 10: Center for Epidemiologic Studies Depression Scale-10; HCHS/SOL: Hispanic Community Health Study/Study of Latinos; HS: High School;

After an average of 6.0 years of follow-up, the age-standardized cumulative incidence of LVH at V2 was 5.4% (95% CI: 4.9, 6.1). The cumulative incidence of LVH at V2 was higher among participants with elevated symptoms of depression at V1 (cumulative incidence= 10.4%, 95% CI: 8.6, 12.4) compared to those with lower symptoms (cumulative incidence = 5.1%; 95% CI: 4.4, 5.9), p value <0.05, **Figure 1**. Similarly, compared with the lowest trait anxiety tertile (cumulative incidence = 5.2%; 95% CI: 4.3, 6.3), the cumulative incidence of LVH at V2 was higher in the middle trait anxiety tertile (8.5%; 95% CI: 7.0, 10.3) and greatest in the highest trait anxiety tertile (9.6%; 95% CI: 8.1, 11.5), p values <0.05.

**Figure 1.**
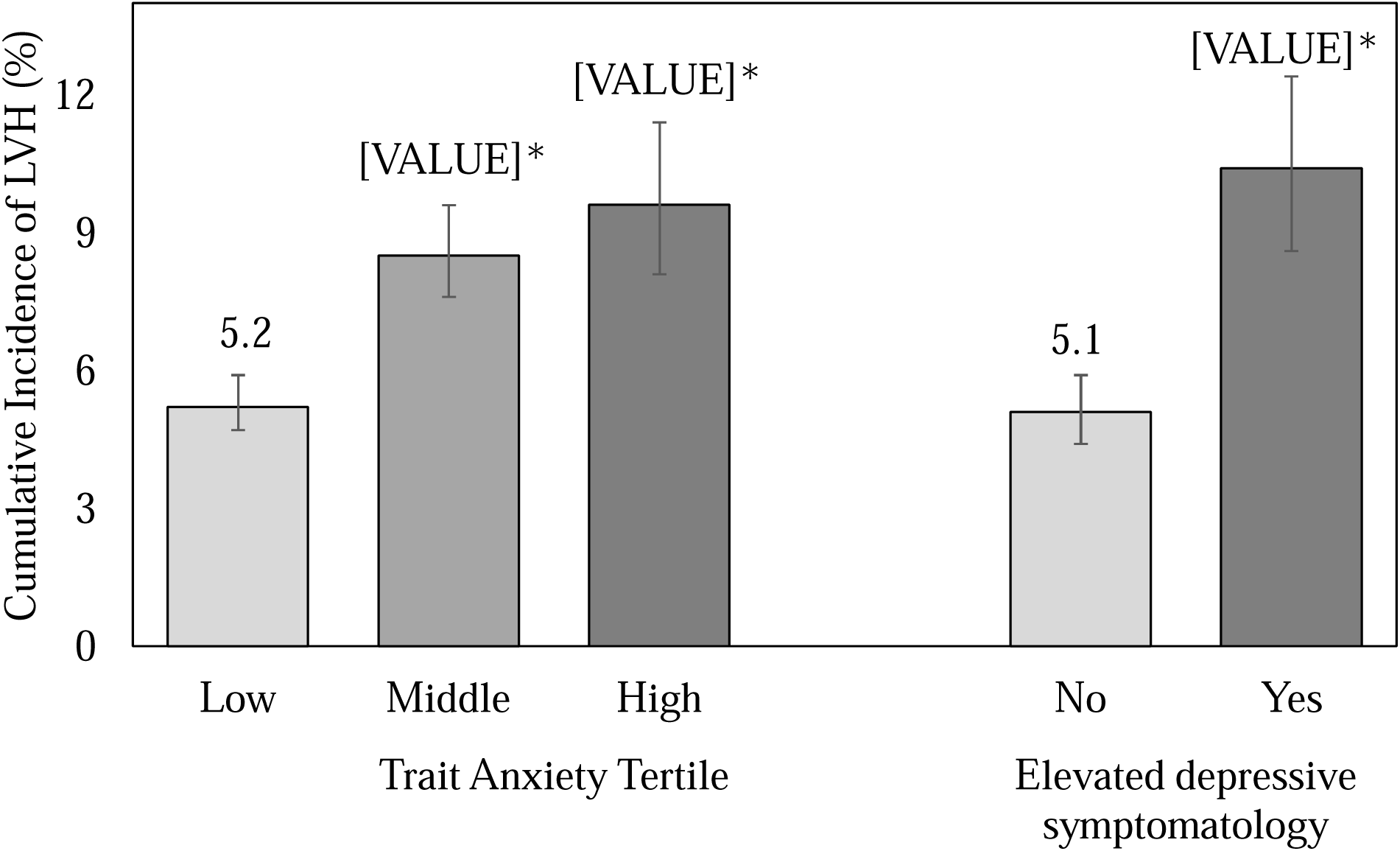
Age standardized incidence of LVH at Visit 2, HCHS/SOL (2014-2017).

In Poisson models adjusted for demographic and socio-economic factors, each standard deviation increment in trait anxiety was not associated with LVH (incidence rate ratio [IRR]: 1.02, 95% CI: 0.92, 1.13), **Table 2**. Likewise, trait anxiety tertile was not associated with LVH. In these same demographic and socio-economic adjusted models, each standard deviation increment in depression symptoms was associated with a 10% greater liklihood of LVH (IRR: 1.10, 95% CI: 1.00, 1.20). Elevated depression symptomatology compared with low depression symptomatology was not significantly associated with LVH. In sensitivity analyses, though the effect size was similar, depression symptomatology was no longer significantly associated with LVH (IRR: 1.12, 95% CI: 1.00, 1.27) after adjusting for potential mediators such as lifestyle/behavioral factors, **Supplemental Table 1**. In another sensitivity analysis, after adjusting for socio-demographic characteristics and hypertension, each standard deviation increment in depression symptomology remained associated with LVH (IDR: 1.09, 1.00, 1.19), **Supplementary Table 2**.

**Table 2:** Multivariable adjusted associations between trait anxiety and depression symptoms in 2008-2011 with LVH in 2014-2017, HCHS/SOL.

|  | Model 1<br>IRR (95% CI) | Model 2<br>IRR (95% CI) |
| --- | --- | --- |
| **Trait Anxiety Score** |  |  |
| 1 SD increment | 1.05 (0.95, 1.15) | 1.02 (0.92, 1.13) |
| Trait Anxiety Tertile |  |  |
| Low (ref) | - | - |
| Middle | 0.97 (0.74, 1.28) | 0.95 (0.72, 1.25) |
| High | 1.05 (0.80, 1.36) | 0.99 (0.75, 1.30) |
| **Depression Symptoms** |  |  |
| 1 SD increment | 1.12* (1.03, 1.22) | 1.10* (1.00, 1.20) |
| Elevated symptoms of depression |  |  |
| No (ref) | - | - |
| Yes | 1.14 (0.92, 1.42) | 1.09 (0.88, 1.35) |

\*Significant at $p < 0.05$
Abbreviations: CI: Confidence interval; HCHS/SOL: Hispanic Community Health Study/Study of Latinos; IRR: Incident rate ratio; LVH: left ventricular hypertrophy; SD: standard deviation
Model 1 is adjusted for age, sex, and Hispanic/Latino background
Model 2 is adjusted age, sex, Hispanic/Latino background, education, income, and health insurance status

In cross-sectional analyses (**Table 3**), each standard deviation increment in the GAD-7 score at V2 was not associated with LVH (OR: 1.04, 95% CI: 0.93, 1.16). Likewise, elevated GAD symptomatology compared with low GAD symptomatology were not associated with LVH (OR: 1.14, 95% CI: 0.83, 1.56) cross-sectionally. Each standard deviation increment in depression symptoms was associated with LVH in demographic-adjusted models (model 1 OR: 1.14, 95% CI: 1.02, 1.27), but not in models adjusted for demographic and socio-economic factors (model 2 OR: 1.09, 95% CI: 0.97, 1.22). Likewise, elevated depression symptomatology was not associated with LVH cross-sectionally. These results remained non-significant in models adjusted for potential mediators (**Supplemental Table 3**). In another sensitivity analysis, after adjusting for socio-demographic characteristics and hypertension, each standard deviation increment in depression symptomology remained associated with LVH (OR: 1.13, 1.00, 1.27) cross-sectionally, **Supplementary Table 4**.

**Table 3:**
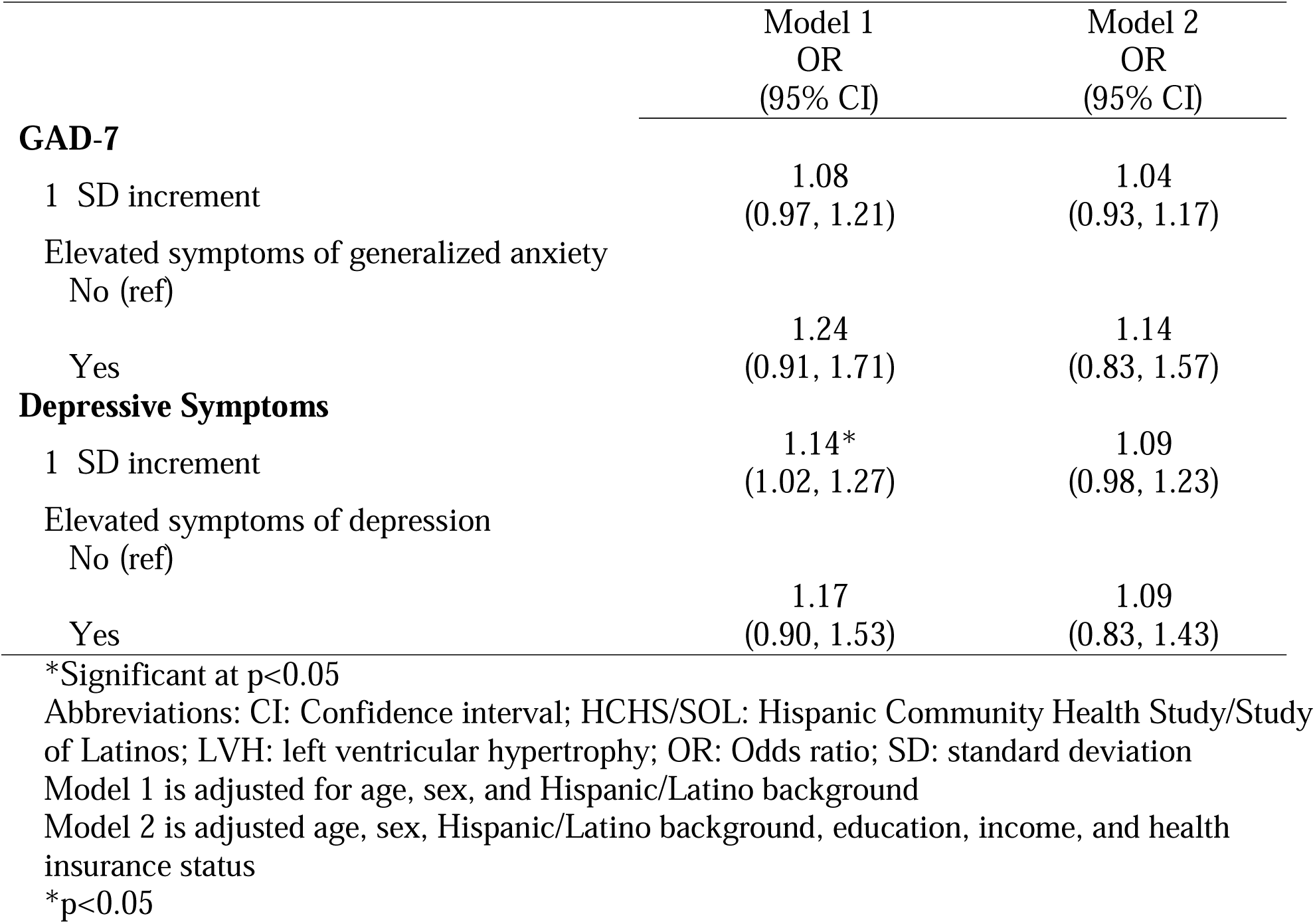
Cross-sectional multivariable adjusted associations between GAD-7, depression symptoms and LVH (2014-2017), HCHS/SOL.

|  | Model 1<br>OR<br>(95% CI) | Model 2<br>OR<br>(95% CI) |
| --- | --- | --- |
| **GAD-7** |  |  |
| 1 SD increment | 1.08<br>(0.97, 1.21) | 1.04<br>(0.93, 1.17) |
| Elevated symptoms of generalized anxiety |  |  |
| No (ref) |  |  |
| Yes | 1.24<br>(0.91, 1.71) | 1.14<br>(0.83, 1.57) |
| **Depressive Symptoms** |  |  |
| 1 SD increment | 1.14*<br>(1.02, 1.27) | 1.09<br>(0.98, 1.23) |
| Elevated symptoms of depression |  |  |
| No (ref) |  |  |
| Yes | 1.17<br>(0.90, 1.53) | 1.09<br>(0.83, 1.43) |

\*Significant at $p < 0.05$
Abbreviations: CI: Confidence interval; HCHS/SOL: Hispanic Community Health Study/Study of Latinos; LVH: left ventricular hypertrophy; OR: Odds ratio; SD: standard deviation
Model 1 is adjusted for age, sex, and Hispanic/Latino background
Model 2 is adjusted age, sex, Hispanic/Latino background, education, income, and health insurance status
\* $p < 0.05$

## DISCUSSION

In a large study of U.S. Hispanic/Latino adults, 5.4% of participants were found to have LVH measured by echocardiography over a six-year period. Participants with higher levels of depression symptomatology or trait anxiety symptomatology at baseline were associated with LVH. However, after adjusting for demographic and socio-economic factors, only depression symptomatology was associated with LVH. These results demonstrate a link between negative affect, and specifically depression symptomatology, with cardiovascular health among Hispanic/Latino adults.

The association between depression and an increased risk of CVD, including sub-clinical disease such as LVH or increased left ventricular mass,^29,46–49^ has been demonstrated in several studies.^25–27,50^ Research in non-Hispanic/Latino populations has found greater endorsement of depression symptomatology to be associated with higher occurrence of LVH or left ventricular alterations among those without hypertension^46–49^ and among those with hypertension,^29,51^ an independent risk factor for LVH.^52^ Cross-sectional studies have found higher levels of depression were associated with greater LVMI^29,46^ and a higher likelihood of LVH,^29,47,48^ independent of blood pressure.^51^ These results were corroborated in the only longitudinal study (to the best of our knowledge) examining the association between depression symptoms and LVH in a cohort of U.S. Black/White adults.^49^ Among female participants in the Coronary Artery Risk Development in Young Adults (CARDIA) Study, sub-threshold or stable depression score on the CES-D 20-item scale was associated with LVH after a 20-year follow-up period.^49^ Our study extends the evidence to U.S. Hispanic/Latino adults, showing elevated depression symptomatology was associated with a risk of developing LVH after a six-year follow-up period, even after adjustment for demographic, socio-economic characteristics, and hypertension in a minimally adjusted model. These results further contribute to the literature, which implicates depression as a strong contributor to adverse cardiovascular outcomes.

Anxiety is associated with the development of hypertension,^23,25,31^ a leading risk factor for LVH.^53^ Among NHW and NHB adults from the National Health and Nutrition Examination Survey I Epidemiologic Follow-up Study, high compared to low levels of anxiety symptoms were associated with an increased risk of incident hypertension among middle-aged NHW adults (45-64 years) and NHB adults aged 25-64 years after a 7-13 follow-up period.^54^ Similarly, research from the HCHS/SOL cohort, one of the first studies to include diverse Hispanic/Latino adults, showed that elevated compared to low anxiety symptoms were associated with a higher incidence of hypertension after a six-year follow-up period.^31^ Anxiety and anxiety disorders have also been linked with an increased risk of CVD,^55,56^ independent of depression and CVD risk factors, such as obesity and metabolic syndrome.^56^ In fact, anxiety symptomatology was associated with a 26% increased risk of coronary heart disease after an average follow-up period of 11 years.^57^ Studies assessing the relationship between negative psychological regulation and LVH are sparse. In one of the few studies conducted among non‐Hispanic/Latino populations, anxiety disorder, and higher anxiety symptomatology were associated with LVH and greater LVMI, a marker of LVH,^58,59^ among hypertensive Turkish adults.^60^ In our study, one of few longitudinal studies to examine the relationship between anxiety and LVH among a cohort of U.S. Hispanic/Latino adults, we did not find a significant association after adjustment for socio-demographic characteristics.

Mechanisms through which anxiety and depression symptomatology can lead to LVH have focused on the relationship between these factors with hypertension. For example, anxiety is associated with increased arterial stiffness,^61,62^ poorer nighttime dipping,^63^ nocturnal and early morning hypertension and diminished circadian rhythm of blood pressure.^64^ Anxiety has also been linked with increased blood pressure variability,^65^ a risk factor for increased LVMI and LVH.^66^ Another proposed pathway between anxiety and the development of hypertension and LVH has been posited from prolonged activation of the hypothalamic-pituitary-adrenal axis and autonomic nervous system as a result of exaggerated neurobiological sensitivity.^67^ Plasma levels of adrenomedullin, which is involved in cardiac remodeling,^68^ are increased with chronic sympathetic nerve excitation^67^ and have been shown to be elevated prior to the development of hypertension^68^ and in individuals with higher LVMI and LVH.^59^ Depressive symptomatology has also been shown to disrupt the hypothalamic-pituitary-adrenal axis.^69^ Higher circulating catecholamines, such as norepinephrine, activate the sympathetic nervous system, resulting in greater cardiovascular load and an increased risk for LVH.^70^ Further, an increase in sympathetic nervous system activity leads to uncontrolled blood pressure, a risk factor for hypertension and subsequent LVH.^27^

Previous research in the HCHS/SOL has found that roughly 40% of the cohort endorsed moderate or high anxiety-depression symptomatology.^71^ Psychological distress, a collective term that includes anxiety and depression symptomatology, was found to be associated with poor cardiovascular health in the HCHS/SOL.^72^ Despite the negative impact of psychological distress on cardiovascular health, primary care does not routinely screen for depression or anxiety.^73,74^ In fact, the U.S. Preventive Services Task Force calls for increased screening of anxiety ^75^ and depression.^76^ Among a cohort of African American adults with hypertension, reduction of stress prevented LVMI progression after a six-month follow-up period.^77^ However, the efficacy of stress reduction on LVMI regression among Hispanic/Latino adults remains unknown. Alternative treatment approaches in LVMI regression and LVH prevention include use of anti-hypertensive agents.^78^ However, prior work in the HCHS/SOL has found that less than 40% of the cohort was treated for hypertension at baseline.^79^ Thus, earlier identification of anxiety or depression symptomatology would help mitigate the downstream pathologic effects caused by negative psychological regulation.

This study is not without limitations. LVH manifests slowly over several years. The average follow-up period in our study was six years. Given the high incidence of hypertension at V2 of the HCHS/SOL,^79^ abnormal blood pressure over a more extended follow-up period may result in the development of LVH among those without a previous diagnosis of LVH. Nonetheless, we still found a significant association between depression symptomatology and LVH. LVH is more accurately assessed by echocardiography than ECG.^80–82^ Therefore the exclusion of individuals with LVH at V1 (measured by ECG) may not fully exclude all participants with LVH at V1 due to lesser sensitivity. However, standardized measurement of ECG abnormalities from the Minnesota code classification and using a central reading center helped minimize potential information bias.^83^ Though cardiac magnetic resonance imaging (MRI) is considered the gold standard for estimating LV mass, it is limited by cost, availability, expertise and certain restrictions, such as the presence of MRI-incompatible devices or metallic remnants.^84,85^ 3D echocardiography approximates MRI in accuracy but is also limited by availability and expertise, making use of 2D echocardiography most practical.^84^ We could not assess the association between symptoms of generalized anxiety disorder and LVH in our main analysis as only trait anxiety was measured at V1. However, we conducted a cross-sectional sensitivity analysis using V2 information and found no significant association between generalized anxiety and LVH. Though we highlight that these two measures may not be comparable as the Spielberger Trait Anxiety Scale measures state and trait non-disorder-specific anxiety while the GAD-7 is used to identify possible cases of GAD and to assess symptom severity. Though we found a significant association between depression symptoms and incident LVH, we may not have been powered to detect differences by anxiety. Scrutiny of the directionality of the beta coefficients for anxiety symptoms in our V2 sensitivity analyses was similar to those of depression symptoms from our primary longitudinal analyses. High stress and low resilience are possible confounders in anxiety and depression symptomatology as well as LVH but we were unable to account for these at V1 and V2. Our study also has important strengths. This study contributes to the sparse literature on the longitudinal relationship between anxiety and depression symptoms with LVH in a cohort of diverse U.S. Hispanic/Latino adults. This study also measured anxiety and depression symptoms using ubiquitous screening tools.^86,87^ Utilizing the CES-D scale and the Spielberger Trait Anxiety Scale is more cost-effective than obtaining a clinical diagnosis. However, as these are not diagnostic tools, clinically diagnosed depression and anxiety possibly may have yielded more robust associations.

In summary, in a population-based sample of U.S. Hispanic/Latino adults, we found higher depression symptomatology was associated with LVH. Our results support the literature showing that negative affect regulation is associated with cardiovascular health. Identifying individuals with adverse psychological regulation and addressing these issues may help mitigate the development of cardiovascular diseases among Hispanic/Latino adults.

## Data Availability

All data produced in the present study are available upon reasonable request to the authors

---

**Supplemental Table 1.** Multivariable adjusted associations between trait anxiety and depression symptoms (2008-2011) with LVH (2014-2017), after adjustment for potential mediators, HCHS/SOL.

|  | IRR (95% CI) |
| --- | --- |
| **Trait Anxiety Score** |  |
| 1 SD increment | 1.00 (0.90, 1.11) |
| Trait Anxiety Tertile |  |
| Tertile 1 (ref) | - |
| Tertile 2 | 0.94 (0.71, 1.24) |
| Tertile 3 | 0.95 (0.72, 1.26) |
| **Depression Symptoms** |  |
| 1 SD increment | 1.06 (0.97, 1.16) |
| Elevated symptoms of depression |  |
| No (ref) | - |
| Yes | 1.02 (0.82, 1.27) |

Models are adjusted for: demographics (age, sex, Hispanic/Latino background), socio-economic status (education, income, health insurance), and potential mediators (alcohol use, smoking status, and diet score) Abbreviations: CI: Confidence interval; HCHS/SOL: Hispanic Community Health Study/Study of Latinos; IRR: Incident rate ratio; LVH: left ventricular hypertrophy; SD: standard deviation

**Supplemental Table 2.** Multivariable adjusted associations between trait anxiety and depression symptoms (2008-2011) with LVH (2014-2017), with adjustment for hypertension, HCHS/SOL.

|  | Incident Rate Ratio<br>(95% Confidence Interval) |
| --- | --- |
| **Trait Anxiety Score** |  |
| 1 SD increment | 1.02 (0.92, 1.12) |
| Trait Anxiety Tertile |  |
| Tertile 1 (ref) | - |
| Tertile 2 | 0.95 (0.72, 1.25) |
| Tertile 3 | 0.99 (0.76, 1.30) |
| **Depression Symptoms** |  |
| 1 SD increment | 1.09* (1.00, 1.19) |
| Elevated symptoms of depression |  |
| No (ref) | - |
| Yes | 1.08 (0.87, 1.33) |

Abbreviations: HCHS/SOL: Hispanic Community Health Study/Study of Latinos; LVH: left ventricular hypertrophy; SD: standard deviation
Adjusted for: age, sex, Hispanic/Latino background, and hypertension
\*p<0.05

**Supplemental Table 3:** Cross-sectional multivariable adjusted associations between GAD-7, depression symptoms and LVH (2014-2017), after adjustment for potential mediators, HCHS/SOL.

|  | Odds Ratio<br>(95% Confidence Interval) |
| --- | --- |
| **GAD-7** |  |
| 1 SD increment | 1.02<br>(0.91, 1.14) |
| Elevated symptoms of generalized anxiety |  |
| No (ref) |  |
| Yes | 1.06<br>(0.77, 1.47) |
| **Depressive Symptoms** |  |
| 1 SD increment | 1.08<br>(0.96, 1.20) |
| Elevated symptoms of depression |  |
| No (ref) |  |
| Yes | 1.04<br>(0.80, 1.37) |

Abbreviations: HCHS/SOL: Hispanic Community Health Study/Study of Latinos; LVH: left ventricular hypertrophy; SD: standard deviation
Adjusted for: age, sex, Hispanic/Latino background, education, income, health insurance, and potential mediators (alcohol use and smoking status)

**Supplemental Table 4:** Cross-sectional multivariable adjusted associations between GAD-7, depression symptoms and LVH (2014-2017), after adjustment for hypertension, HCHS/SOL.

|  | Odds Ratio<br>(95% Confidence Interval) |
| --- | --- |
| **GAD-7** |  |
| 1 SD increment | 1.06<br>(0.95, 1.19) |
| Elevated symptoms of generalized anxiety |  |
| No (ref) |  |
| Yes | 1.18<br>(0.85, 1.63) |
| **Depressive Symptoms** |  |
| 1 SD increment | 1.13*<br>(1.00, 1.27) |
| Elevated symptoms of depression |  |
| No (ref) |  |
| Yes | 1.20<br>(0.91, 1.58) |
| Abbreviations: GAD: Generalized anxiety disorder; HCHS/SOL: Hispanic Community Health Study/Study of Latinos; LVH: left ventricular hypertrophy; SD: standard deviation |  |
| Models are adjusted for age, sex, Hispanic/Latino background and hypertension |  |
| *p<0.05 |  |

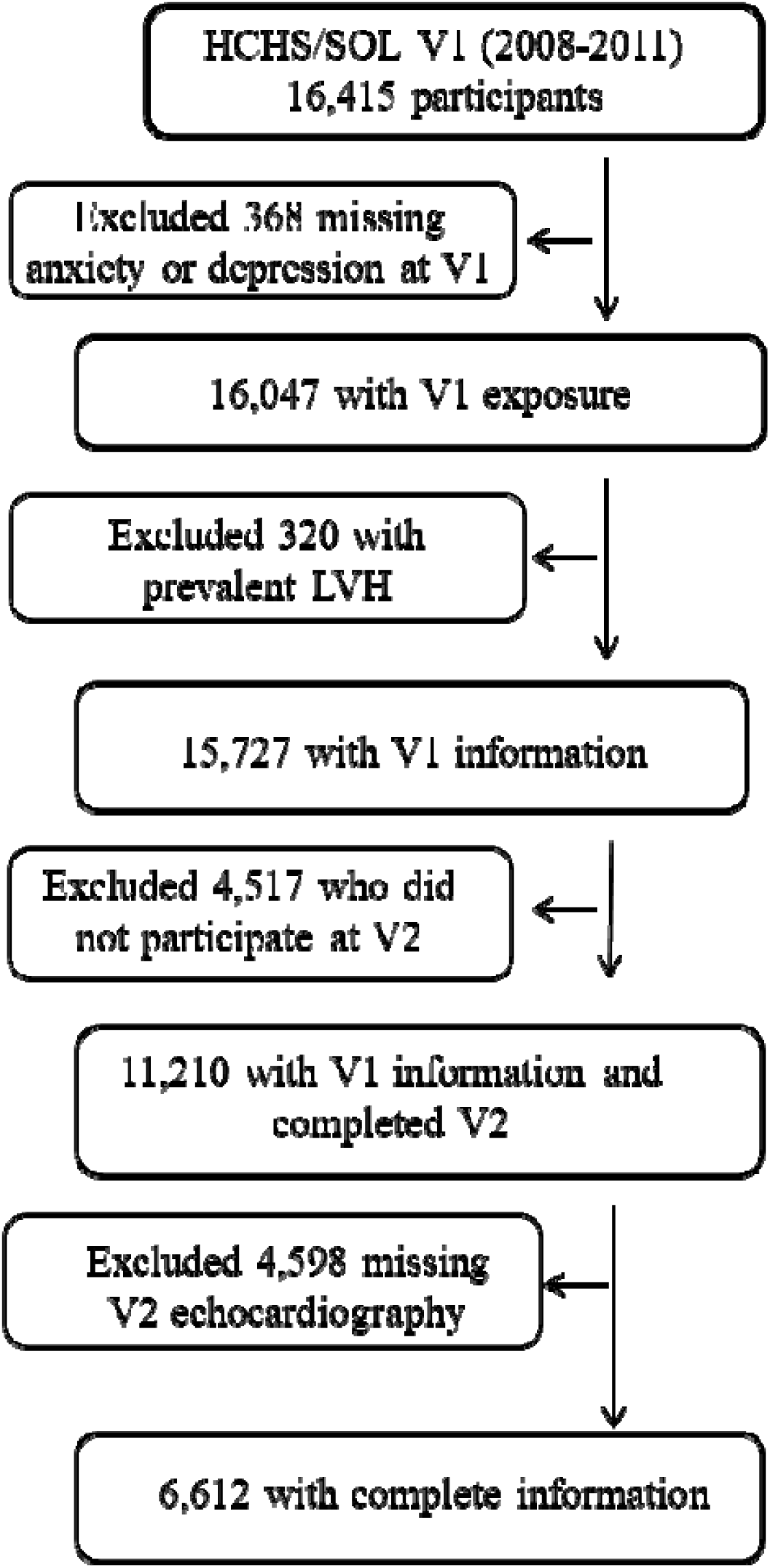

